# Patient Preferences to Undergo Low-Value CT Coronary Angiography in the Emergency Department

**DOI:** 10.1101/19008391

**Authors:** Jessica L. Winkels, Chelsea Morrow Smith, Rahul Iyengar, Arjun P. Meka, Jonathan D. Porath, William J. Meurer

## Abstract

**Background:** Low-value diagnostic testing adds billions of dollars to the cost of health care in the US annually. Addressing patient preference for these tests is one possible strategy to limit overuse. In previous work, we showed that patient preference for testing can be influenced by test benefit, risk, and financial measures. Our objective was to examine the effect of these variables in another clinical scenario involving chest pain.

**Methods:** In this cross-sectional survey of patients at the University of Michigan Emergency Department (ED), participants were given a hypothetical scenario involving an ED visit for chest pain, along with information regarding potential benefit (detecting a life-threatening condition; 0.1 or 1%) and risk (developing cancer; 0.1 or 1%) of CTCA, as well as an incentive of $0 or $100 to forego testing. Values for risk, benefit, and financial incentive varied across participants. Our primary outcome was patient preference to undergo testing. We also obtained demographic and numeracy information. Then, we used logistic regression to estimate odds ratios, adjusting for multiple potential confounders. Our sample size was designed to find at least 300 events (test acceptance) to allow for up to 30 covariates in fully adjusted models. We had 85-90% power to detect a 10% absolute difference in testing rate across groups, assuming a 95% significance level.

**Results:** 913 patients were surveyed. A $100 financial incentive (adjusted OR [AOR] 0.57; 95% Confidence Interval [CI] 0.42-0.78) and increased test risk (AOR 0.61; 95% CI 0.44-0.84) both significantly decreased test acceptance in fully adjusted models, whereas increased test benefit significantly increased test acceptance (AOR 2.45; 95% CI 1.79-3.36).

**Conclusions:** Offering a financial incentive deterred patients from accepting testing despite varying levels of risk and benefit. In the context of previous work, we provide preliminary evidence supporting that financial interventions may impact patient preference more than test risk and benefit.

## Introduction

Overuse of diagnostic testing in low-risk patients contributes substantially to the rising cost of health care. One study estimated that the total annual financial burden of defensive medicine in the United States exceeded $46 billion in 2008 alone, and it is likely that this figure has continued to rise in proportion with the overall cost of health care.^1^ Furthermore, patients who undergo unnecessary diagnostic tests are still subject to the risks of testing when potential benefit is limited. Computed Tomography Coronary Angiography (CTCA) is a diagnostic test has clinical utility in determining the need for more invasive procedures in patients with chest pain and has a high negative predictive value for ruling out anginal chest pain.^2^ While it is useful for certain patients, CTCA also involves radiation exposure that has been associated with an increased cancer risk.^3^

Addressing overuse of low-value testing can be difficult, in part because the term “low-value” is often subjectively defined. Although there are clinical decision tools available for providers to risk stratify patients, the potential risk and benefit of testing varies considerably among patients.^4^ Initiating discussions with patients regarding the benefits and risks of low-value testing to elicit patient preference has emerged as a potential solution to decrease the use of unnecessary diagnostic testing in the emergency department. There is also evidence to support that patients may be responsive to financial information while making health care decisions, although this continues to be debated and has yet been be fully described.^5-9^

Our previous work on this topic demonstrated that increased financial cost to patients significantly decreased their desire for low-value diagnostic testing with CTCA and Computed Tomography (CT) head scans in hypothetical scenarios involving chest pain and minor traumatic brain injury (TBI), respectively.^7, 9^ A subsequent study focused solely on patient preferences for CT head in minor TBI showed that a financial incentive for patients to forego low-value diagnostic testing also significantly reduced patient preference for this test.^6^ While the influence of test benefit and risk on patient decision-making was of mixed statistical significance in our original work, these variables both achieved statistical significance in our CT head/minor TBI study.^6, 9^

In this study, we investigated the effect of a direct financial incentive to forego low-value diagnostic testing on patient preference to pursue diagnostic CTCA in a hypothetical scenario involving low-risk chest pain in the context of varying levels of associated test benefit and risk. Our aim in this study was to obtain further data to better characterize the effect of a financial incentive – as well as varying levels of test benefit and risk – on patient preference across another clinical scenario involving a different diagnostic test. We hypothesized that our results would be comparable to our follow-up study involving minor TBI in that a financial incentive, increased test risk, and decreased test benefit would all significantly deter patients from accepting testing with low-value CTCA.

## Methods

### Overview

This study is a cross-sectional survey of a convenience sample of adult patients from the University of Michigan Emergency Department (ED) designed to examine the potential effect of financial incentive, test benefit, and test risk on patient preference for low-value diagnostic testing with CTCA in the setting of chest pain. The current study reports a different outcome based on previously reported methodology.^6^

### Study Design

Study participants were presented with a hypothetical scenario in which they presented to the ED with low-risk chest pain, for which further testing with CTCA would not be clinically indicated. All patients were assigned values for potential test benefit (percent chance that a CTCA would detect a life-threatening acute process; either 0.1 or 1%) that would classify them as having a HEART score in the low risk category.^10^

The full study scenario is available in Appendix A. Consent was obtained, and the scenario script was read aloud to all participants to minimize any issues they might have with reading, seeing, or understanding the scenario. Participants then were assigned a value for test benefit (detecting a life-threatening medical condition; either 0.1% or 1%), test risk (the chance of developing cancer due to radiation within 10 years; either 0.1% or 1%), and financial incentive (a cash payment of $0 or $100 from an insurance company to forego CTCA) prior to being asked for their preference for or against testing. Values for test risk and benefit were represented as percentages, ratios, and visual depictions (Appendix A) to facilitate participants’ understanding.^11, 12^ As in previous work, these values for benefit, risk, and financial incentive were chosen because they were thought to encompass the most interesting zone of variation in patient preference in a preliminary study performed by the authors.^7^ The risk values were also considered plausible benefit and risk probabilities associated with diagnostic CTCA.^13^

### Setting and Population

Our study used a convenience sample of 913 adult patients in the University of Michigan ED who were not presenting with chest pain, recent head trauma, or altered mental status. No patients in resuscitation bays or with contact precautions were approached. Participation was voluntary, and participants were not offered financial compensation for participating.

### Human Subjects Protection

The University of Michigan Institutional Review Board reviewed this study and determined it to be exempt survey research.

### Primary Outcomes and Variables

Our primary outcome was the percentage of patients accepting diagnostic testing with CTCA given the three major predictive variables: test benefit, test risk, and financial incentive. This combination of variables resulted in eight possible subgroups. We obtained de-identified demographic information and relevant past medical history to assess for potential confounders, which were specified in advance. Confounders included age, gender, marital status, educational status, race, ethnicity, income, prior medical training or employment, self-reported overall health, and past medical history of cancer, hypertension, diabetes, atrial fibrillation, myocardial infarction, or head trauma requiring a hospital visit. Numeracy was evaluated by a previously validated assessment, and participants were classified as having low, medium, or high numeracy accordingly.^14^

#### Data Collection

We used Qualtrics for survey administration and data collection, and we used SPSS (Armonk, NY Version 25) for data analysis. All participant responses in which the primary outcome was obtained were incorporated into the analysis. We compared the unadjusted proportion of participants who chose to receive a CTCA for each combination of values for test benefit, test risk, and financial incentive.

### Sample Size

We followed methodology that was previously reported in our 2018 and 2019 work analyzing the effect of an additional copayment or financial incentive for a diagnostic test, respectively.^6, 9^ Our sample size of 913 conferred approximately 85-90% power to detect a 10% absolute change in the proportion of subjects agreeing to testing from a baseline test acceptance rate of 50% at a 95% level of significance.

### Data Analysis

We performed a series of nested multivariable logistic regression models to obtain the odds that participants would agree to undergo a CTCA, given the variable combinations as noted above. We adjusted for four sets of variables in the models, and all variables were specified in advance. Variables were ordered so that those predicted to be more influential would be included in earlier models. The fully adjusted model was limited to at most 30 variables in order to not exceed 10 outcome events per predictor. Model 1 adjusted for the benefit, risk, and financial incentive associated with testing. Model 2 additionally adjusted for income, education level, and numeracy. Model 3 additionally adjusted for age, gender, race, ethnicity, and previous healthcare training/employment. Finally, Model 4 additionally adjusted for self-reported overall health and a medical history of cancer, hypertension, diabetes, atrial fibrillation, myocardial infarction, or head trauma requiring a hospital visit. Model fit was evaluated by the Hosmer and Lemeshow Goodness of Fit Statistic with a p value of >0.05 indicating adequate fit. The deidentified dataset and model output (including all parameter estimates for the fully adjusted models, and goodness of fit statistics) is posted in the University of Michigan Institutional Data Repository (https://doi.org/10.7302/pnmm-4v40).

## Results

913 patients met inclusion criteria and completed the primary outcome by submitting their preference regarding undergoing diagnostic testing with CTCA. These 913 participants’ results were incorporated into the analysis. The median participant age was 46 years (interquartile range 30-60) with an absolute range of 18-92 years. Further demographic and medical characteristics are displayed in Table 1.

**Table 1:** Characteristics of Study Participants (N = 913)

| Characteristic | % (n) |
| --- | --- |
| Age, years |  |
| 18-25 | 16% (146) |
| 26-40 | 23.1% (211) |
| 41-55 | 25.6% (234) |
| 56-65 | 15.0% (137) |
| 66-75 | 10.7% (98) |
| > 76 | 5.1% (47) |
| Unreported | 4.4% (40) |
| Sex |  |
| Male | 39.6% (362) |
| Female | 56.1% (512) |
| Other/Transgender | 0.1% (1) |
| Unreported | 4.1% (38) |
| Marital status |  |
| Married | 49.8% (455) |
| Divorced | 7.6% (69) |
| Single/never married | 32.0% (292) |
| Separated | 1.2% (11) |
| Widowed | 5.0% (46) |
| Unreported | 4.4% (40) |
| Highest level of education |  |
| Some high school | 3.9% (36) |
| High school graduate | 15.4% (141) |
| Some college | 31.5% (288) |
| College graduate | 26.4% (241) |
| Post-graduate | 16.1% (147) |
| Unreported | 6.6% (60) |
| Works in healthcare | 24.5% (224) |
| Hispanic | 5.3% (48) |
| Race |  |
| American Indian/Alaska Native | 0.5% (5) |
| African American | 12.0% (110) |
| Caucasian | 77.1% (704) |
| Asian | 2.1% (19) |
| Native Hawaiian/Pacific Islander | 0.2% (2) |
| Other | 2.0% (18) |
| Prefer not to disclose/Unreported | 6.0% (55) |
| History of cancer | 13.2% (120) |
| History of diabetes | 15.1% (137) |
| History of hypertension | 29.2% (264) |
| History of atrial fibrillation | 7.7% (70) |
| History of heart attack | 5.0% (45) |
| History of head injury requiring ED visit | 20.5% (184) |
| Self-reported overall health |  |
| Excellent | 10.6% (97) |
| Very good | 26.2% (239) |
| Good | 28.3% (258) |
| Fair | 18.4% (168) |
| Poor | 9.1% (83) |
| Unreported | 7.5% (68) |
| Household income level |  |
| Less than \$10,000 | 5.1% (47) |
| \$10,000 – \$14,999 | 2.8% (26) |
| \$15,000 – \$24,999 | 3.6% (33) |
| \$25,000 – \$34,999 | 7.3% (67) |
| \$35,000 – \$49,999 | 6.0% (55) |
| \$50,000 – \$74,999 | 9.7% (89) |
| \$75,000 – \$99,999 | 7.4% (68) |
| \$100,000 – \$149,999 | 10.0% (91) |
| \$150,000 – \$199,999 | 3.2% (29) |
| \$200,000 or more | 5.4% (49) |
| Unreported/Prefer not to disclose | 39.3% (359) |

Patients agreed to undergo CTCA in 62.8% of scenarios (573 out of 913). Unadjusted patient preferences in each of the eight subgroups, which account for all possible combinations of the three predictor variables, are shown in Table 2. In the unadjusted models, we found that increased test risk and a financial incentive to forego testing resulted in a statistically significant decrease in odds of test acceptance, whereas increased test benefit increased patient preference for testing (Table 3). Each of the adjusted models also demonstrated this relationship (Table 4), suggesting that none of the variables incorporated into the adjusted models were confounders influencing the observed effect of the major predictive variables on test acceptance.

**Table 2:** Patient Preferences by Subgroup.

| Incentive = \$0 | | | |
| --- | --- | --- | --- |
|  | Risk |  |  |
|  |  | 0.1% | 1% |
| Benefit | 0.1% | Accept Test: 66.1%<br>(74 of 112) | Accept Test: 48.7%<br>(58 of 119) |
|  | 1% | Accept Test: 86.6%<br>(84 of 97) | Accept Test: 72.5%<br>(95 of 131) |

| Incentive = \$100 | | | |
| --- | --- | --- | --- |
|  | Risk |  |  |
|  |  | 0.1% | 1% |
| Benefit | 0.1% | Accept Test: 53.9%<br>(69 of 128) | Accept Test: 42.7%<br>(41 of 96) |
|  | 1% | Accept Test: 67.2%<br>(80 of 119) | Accept Test: 64.9%<br>(72 of 111) |

**Table 3:** Unadjusted Patient Preferences^*^.

|  | % (n) accepting test |
| --- | --- |
| Benefit |  |
| 0.1% (ref) | 53.1% (242) |
| 1% | 72.3% (331) |
| OR (CI 95%) | 2.287 (1.738-3.008) |
| Risk |  |
| 0.1% (ref) | 67.3% (307) |
| 1% | 58.2% (226) |
| OR (CI 95%) | 0.673 (0.514-0.882) |
| Incentive |  |
| \$0 (ref) | 67.8% (311) |
| \$100 | 57.7% (262) |
| OR (CI 95%) | 0.647 (0.494-0.847) |
| Total | 62.8% (573 of 913) |

\* OR = odds ratio; CI = confidence interval All odds ratios are unadjusted

**Table 4:** Nested Logistic Regression Model^*^.

|  | Model 1<br>AOR (95% CI) | Model 2<br>AOR (95% CI) | Model 3<br>AOR (95% CI) | Model 4<br>AOR (95% CI) |
| --- | --- | --- | --- | --- |
| Benefit<br>(1% vs. 0.1%) | 2.42 (1.83-3.20) | 2.47 (1.82-3.35) | 2.42 (1.78-3.29) | 2.45 (1.79-3.36) |
| Risk<br>(1% vs. 0.1%) | 0.60 (0.45-0.79) | 0.61 (0.45-0.83) | 0.60 (0.44-0.82) | 0.61 (0.44-0.84) |
| Incentive<br>(\$100 vs \$0) | 0.60 (0.45-0.79) | 0.57 (0.42-0.77) | 0.58 (0.43-0.79) | 0.57 (0.42-0.78) |

\* Model 1 adjusts for benefit, risk, and incentive associated with testing. Model 2 additionally adjusts for income, education level, and numeracy. Model 3 additionally adjusts for age, gender, race, ethnicity, and previous healthcare training or employment. Model 4 additionally adjusts for self-reported overall health and a medical history of cancer, hypertension, diabetes, atrial fibrillation, myocardial infarction, and head trauma requiring hospital visit. Hosmer and Lemeshow Goodness of Fit p-value ranged from 0.9 to 0.3, indicating that model fit was adequate. AOR = adjusted odds ratio; CI = confidence interval.

The fully adjusted models (Table 4) indicated that the odds of patients accepting a CTCA was significantly lower when patients were offered a $100 financial incentive to forego testing, as compared to when no incentive was offered (adjusted OR [AOR] 0.57; 95% Confidence Interval [CI] 0.42-0.78). Increasing test benefit from 0.1% to 1% resulted in a significant increase in odds of test acceptance (AOR 2.45; 95% CI 1.79-3.36), and increasing test risk from 0.1% to 1% was associated with a significant decrease in odds of test acceptance (AOR 0.61; 95% CI 0.44-0.84).

## Discussion

This study examined the effect of test benefit, test risk, and a financial incentive on patient preference for pursuing low-value CTCA for chest pain in the ED. We found that decreased test benefit, increased test risk, and offering a financial incentive decreased patient preference for testing in a statistically significant manner. These findings supplement our prior work by providing further evidence that a financial intervention targeted at reducing patient preference for low-value testing may be effective in influencing patients’ decision-making. A financial incentive thereby may help to decrease costs attributable to unnecessary diagnostic testing in the ED. Additionally, our data provides further evidence to suggest that discussing benefits and risks of diagnostic testing with patients can impact their preferences regarding low-value tests, even when values for benefit and risk are low.

These results are particularly interesting in the context of our prior study in 2018, which evaluated the impact of benefit, risk, and out-of-pocket cost on patient preference for low-value CTCA in a similar hypothetical low-risk chest pain scenario.^9^ In the current study, there was a 19.2% drop in test acceptance (72.3% to 53.1%) with decreased test benefit, a 9.1% drop (67.3% to 58.2%) with increased risk, and a 10.1% drop (67.8% to 57.7%) with a financial incentive. In the 2018 work, we observed a similar trend in the data – there was a 9.4% drop (74.3% to 65.0%) in test acceptance with decreased benefit, a 1.5% drop (70.4% to 68.9%) with increased risk, and a 22% drop (80.8% to 58.7%) with increased cost to the patient, although only the effects of benefit and cost reached statistical significance. Additionally, the results of our current study were congruent with our more recent study that analyzed the effect of test benefit, test risk, and financial incentive on patient preference for low-value diagnostic testing with CT head in a minor TBI scenario.^6^ This supports that the relationship between these variables may be observed across multiple clinical situations.

Overall, the consistent effect of a financial intervention across all three studies provides preliminary evidence to suggest that a financial strategy may be effective in modulating patients’ preferences regarding low-value diagnostic testing in the ED. Furthermore, while varying test risk and benefit did influence patients’ odds of test acceptance, these variables were not statistically significant in all three studies. This suggests that a financial approach may be more effective in influencing patient preferences than discussing the risks and benefits of testing alone. Further study will be required to better describe the relationship between these variables and patient preferences, as well as to comment on the generalizability of our findings in clinical scenarios beyond low-risk chest pain and minor TBI.

### Limitations

There are several limitations that should be considered when interpreting our results. First, we surveyed ED patients using a hypothetical clinical scenario that was intentionally different from their presenting chief complaint. Patients experiencing chest pain in the ED may be influenced differently by our predictor variables. Additionally, the hypothetical scenario in our study provided numeric representations of benefit and risk, whereas an individual patient’s actual benefit and risk for a particular test may not be easily quantifiable in this way. True risk of CTCA may be lower than the percentages assigned in these scenarios, but 0.1% and 1% were selected to maintain symmetry with values for potential benefit. Our study also cannot account for the varying levels of medical knowledge or past exposure to health care that may have affected patients’ decision-making, even though participants were instructed to only consider the numeric values for benefit and risk. Furthermore, as with prior studies, these hypothetical scenarios required patients to consider the benefit of detecting an acute medical condition against the risk of an adverse outcome far in the future – different time of onset for benefit and risk may have also influenced patients’ decisions. Finally, while we have now shown that a financial incentive is associated with a statistically significant decrease in test acceptance in two distinct clinical scenarios, this may not be generalizable outside the context of a low-risk, low-value test. Patients may also make decisions differently if a test is designed to detect a medical condition that is not life-threatening.

### Conclusions

This cross-sectional survey of patients in the ED suggests that a financial incentive may be effective in dissuading patients from accepting low-value testing with CTCA in the context of low-risk chest pain. Given that this effect was also observed in a separate clinical scenario in our prior study with similar methodology, it is possible that offering a financial incentive may be an effective deterrent against patient preference for low-value testing across several distinct clinical scenarios. Lower test benefit and higher test risk also significantly decreased patient preference for testing in this study; however, these variables have not consistently emerged as statistically significant factors in our previous work, whereas financial interventions have been statistically significant throughout. Taken together, the findings in these studies suggest that a financial approach may be more effective in influencing patient preference for low-value diagnostic testing than discussion of test benefit and risk alone. Further study is needed to better characterize the effect of each of these factors on patient preferences in additional clinical scenarios and in a non-hypothetical setting.

## Data Availability

The deidentified dataset and model output (including all parameter estimates for the fully adjusted models, and goodness of fit statistics) is posted in the University of Michigan Institutional Data Repository (https://doi.org/10.7302/pnmm-4v40).

